# COVID-19 Vaccination Induces Cross-Reactive Dengue Antibodies with Altered Isotype Profiles and In Vitro ADE

**DOI:** 10.1101/2025.05.23.25328123

**Authors:** Sebastian Reinig, Chin Kuo, Sheng-Yu Huang, Kuei-Ching Hsiung, Po-Kai Chen, Etsuro Ito, Ing-Kit Lee, Ching-Yen Tsai, Shu-Min Lin, Shin-Ru Shih

## Abstract

Dengue virus is a mosquito transmitted flavivirus that cause inapparent or mild disease but can also cause severe hemorrhagic fever in humans. Cross-reactive IgG antibodies can cause under specific conditions antibody dependent enhancement (ADE) of the infection via Fc-receptor interaction. Antibodies from SARS-CoV-2 (anti-S) causing COVID-19 disease can also cross-react against the dengue envelope (anti-E). We therefore investigated the antibody profile of these cross-reactive antibodies and their ability to induce ADE. We found that the cross-reactive anti-E in COVID-19 vaccinated are dominated by IgM/A while, anti-S from vaccinated or anti-E from dengue infected patients have an IgG1 dominated antibody profile. The low-level anti-E IgG from COVID-19 vaccinated are dominated by low Fc-affinity IgG2/4 subclass. These antibodies were able to induce in vitro ADE stronger than from Dengue infected for the 1^st^,2^nd^ dose of the vaccine or later omicron variant booster. This could partially be explained that ADE was inhibited by the complement system in dengue infected but not in COVID-19 vaccinated.

## Introduction

Dengue virus (DENV) infection poses a significant public health challenge in tropical and subtropical regions, causing over 400 million cases and approximately 20,000 deaths annually. This flavivirus is transmitted by mosquito vectors. While most infections are mild or asymptomatic, a small proportion can progress to severe conditions, such as dengue hemorrhagic fever or dengue shock syndrome^1^. Although adaptive immunity typically provides protection, a secondary dengue infection can result in more severe disease in some individuals. This is believed to be due to antibody-dependent enhancement (ADE). In ADE, IgG antibodies bind to the dengue virus, promoting its entry into mononuclear immune cells through Fc receptors and potentially intensifying macrophage inflammation^2^. The structural envelope (E-protein) of the dengue virus is known to cross-react with antibodies elicited by other viruses, particularly other flaviviruses^3^. It was also demonstrated that antibodies from the immune response against SARS-CoV-2 can cross-react against DENV2 envelope^4^ or for more recent variants also against DENV3 ^5^. In SARS-CoV-2 vaccination, it is recognized that mRNA and protein subunit vaccines induce a gradual shift in antibody subclasses, transitioning from high Fc-receptor affinity IgG1/IgG3 to low Fc-receptor affinity IgG2/IgG4 over time^6–8^. Previous studies have demonstrated that IgG2/IgG4 antibodies, despite sharing the same antigen-binding Fab region, exhibit lower Fc-receptor affinity and reduced maximum in vitro ADE capacity compared to other subclasses^9^. Additionally, an elevated ratio of anti-E IgG1 to IgG2 antibodies is linked to increased disease severity^10^. It is so far unclear whether this affects the isotype distribution of cross-reactive antibodies and their potential to induce ADE. To investigate, we compared the isotypes of anti-dengue antibodies in individuals vaccinated against COVID-19 and those who recovered from dengue. Additionally, we evaluated the cross-reactivity of these antibodies and their capacity to induce ADE.

## Material and Methods

### Sample collection

A total of 271 serum or plasma samples were collected from individuals before vaccination and after the first, second, and third doses of the following vaccines: the adenovector vaccine ChAdOx1 nCoV-19 (AstraZeneca, AZ), the protein subunit vaccine MVC-COV1901 (Medigen), or the mRNA vaccines mRNA-1273 (Moderna) and BNT162b2 (BioNTech/Pfizer) (**Table 1**). Furthermore, 21 samples were collected from individuals which received omicron targeted-booster: Moderna (XBB), Novavax (XBB) and Moderna (Ba.4.5) (**Supplemental table 1**). Additionally, 24 samples from dengue convalescent individuals were collected at Kaohsiung Chang Gung Memorial Hospital or Kaohsiung Municipal Fengshan Hospital between February 10 and December 7, 2023, following confirmation of dengue infection by PCR or ELISA through the hospital’s clinical laboratory **(Supplementary Table 2)**. Serotypes were determined by serum neutralization assay and could identify the serotype in 13 of the 24 samples: 10 were identified as dengue virus serotype 1 (DENV-1), 2 as serotype 2 (DENV-2), and 1 sample showed a mixed infection with both DENV-1 and DENV-2. The samples from Taiwan were collected from individuals at Chang Gung Memorial Hospital and Chang Gung University (ethics approval number: 202001041A3C). Negative human serum samples from the United States were purchased from Access Biologicals (**Supplemental table 3**). These samples were collected as per protocol SDP-003, Human Biological Specimens Collection, dated September 22, 2017, and the qualifications of the principal investigator (Robert Pyrtle, M.D.) were reviewed and approved by the Diagnostics Investigational Review Board (Cummaquid, MA, USA).

**Table I.**
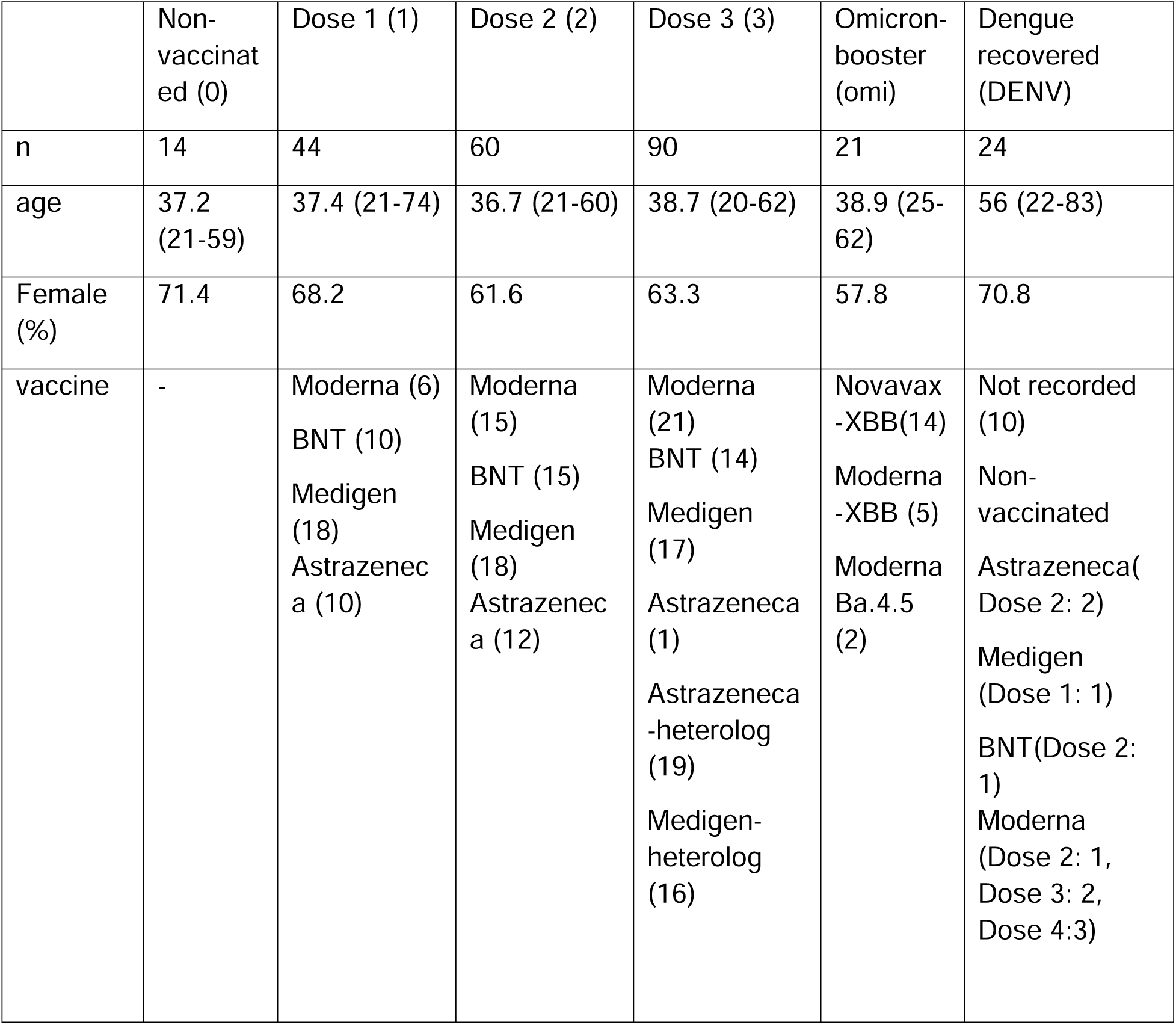
Provide the age and sample collection timing (in days post-infection or symptom onset) for each group, reported as the mean, minimum, and maximum values.

|  | Non-vaccinated (0) | Dose 1 (1) | Dose 2 (2) | Dose 3 (3) | Omicron-booster (omi) | Dengue recovered (DENV) |
| --- | --- | --- | --- | --- | --- | --- |
| n | 14 | 44 | 60 | 90 | 21 | 24 |
| age | 37.2 (21-59) | 37.4 (21-74) | 36.7 (21-60) | 38.7 (20-62) | 38.9 (25-62) | 56 (22-83) |
| Female (%) | 71.4 | 68.2 | 61.6 | 63.3 | 57.8 | 70.8 |
| vaccine | - | Moderna (6)<br>BNT (10)<br>Medigen (18)<br>Astrazeneca (10) | Moderna (15)<br>BNT (15)<br>Medigen (18)<br>Astrazeneca (12) | Moderna (21)<br>BNT (14)<br>Medigen (17)<br>Astrazeneca (1)<br>Astrazeneca-heterolog (19)<br>Medigen-heterolog (16) | Novavax-XBB(14)<br>Moderna-XBB (5)<br>Moderna Ba.4.5 (2) | Not recorded (10)<br>Non-vaccinated<br>Astrazeneca(Dose 2: 2)<br>Medigen (Dose 1: 1)<br>BNT(Dose 2: 1)<br>Moderna (Dose 2: 1, Dose 3: 2, Dose 4:3) |

### Dengue Virus Serum Neutralization Assay

The dengue virus serum neutralization assay was performed using Vero cells in a 96-well format to evaluate serum neutralizing titers against dengue virus isolates. Falcon 96-well cell culture plates were used for cell seeding, while GeneDirex 96-well plates were designated for serum dilution and incubation. Prior to use, all serum samples were heat-inactivated at 56□°C for 30 minutes.

Vero cells were cultured in Minimum Essential Medium (MEM) supplemented with 10% fetal bovine serum (FBS), 2% penicillin-streptomycin (P/S), 1% non-essential amino acids (NEAA), and 1% sodium pyruvate. For virus dilution and infection, a modified medium consisting of MEM with 2% FBS, 2% P/S, 1% NEAA, and 1% sodium pyruvate was used. The overlay medium was prepared by supplementing this modified medium with 1% methylcellulose.

To initiate the assay, Vero cells were seeded at a density of 2.5 × 10⁴ cells in 100 µL per well and incubated overnight at 37□°C in a 5% CO₂ atmosphere. On the following day, serum samples were first diluted 1:10 with the modified diluent medium and then subjected to two-fold serial dilutions ranging from 1:20 to 1:20,480, with 40 µL per dilution well in duplicate. Virus stocks were prepared to achieve approximately 60 foci per well, based on prior back titration results. Equal volumes (40 µL) of diluted virus were mixed with the diluted serum samples, and the mixtures were incubated at 37□°C for 30 minutes. Concurrently, the culture medium was removed from the Vero plates. After incubation, 30 µL of each virus-serum mixture was transferred into the corresponding wells containing Vero cells and incubated at 37□°C for 1 hour to allow virus adsorption, with gentle rocking of the plates after 30 minutes. Following adsorption, 150 µL of pre-warmed overlay medium was added to each well, and the plates were incubated at 37□°C in 5% CO₂ for four days.

On day six, the overlay medium was aspirated, and the wells were gently washed twice with 180 µL of 1× phosphate-buffered saline (PBS). Cells were then fixed with 100 µL of 80% methanol for 10 minutes at room temperature. Plates were either immediately processed for immunostaining or stored at −80□°C for subsequent analysis.

### Immunostaining and Focus Enumeration

Following methanol fixation, the 96-well plates were air-dried and washed twice with 180□μL of phosphate-buffered saline (PBS), followed by gentle blotting to remove residual liquid. To block nonspecific binding, 180□μL of PBS containing 5% non-fat dry milk was added to each well and incubated at room temperature for 30 minutes. The blocking buffer was then removed, and 100□μL of primary antibody solution (1:2000 dilution of Anti-Dengue Virus Complex Antibody, MAB8075) was added to each well. Plates were incubated at room temperature for 2–3 hours with gentle shaking at 30□rpm.

After primary antibody incubation, the wells were washed four times with 180□μL of PBST (PBS containing 0.05% Tween-20), followed by blot drying. Secondary antibody solution (100□μL per well, 1:3000 dilution of Anti-Mouse IgG (gamma) Antibody, Cat. #5220-0460) was then added, and plates were incubated at room temperature for 1 hour with shaking at 30□rpm. Wells were subsequently washed six times with PBST and air-dried.

To visualize the foci, 60□μL of TrueBlue™ Peroxidase Substrate (SeraCare, Cat. #5510-0030) was added to each well and incubated for 20 minutes in the dark. The substrate was then removed, and the plates were air-dried. The number of immunostained foci was quantified using an automated ImmunoSpot analyzer (AID), and viral titers were calculated accordingly.

### Antibody isotype enzyme-linked immunosorbent assay

The full spike trimer D614G or the envelope protein from dengue virus serotype 2 (Sino Biological, Beijing, China) was immobilized at 0.2 mg/mL per well in phosphate-buffered saline (PBS) on a 96-well flat-bottom polystyrene microplate overnight. A blocking solution of 2.5% bovine serum albumin (BSA) in PBST (PBS with 0.05% Tween 20) was applied. All samples were diluted in PBST under identical conditions: 1:200 for anti-E IgG subclasses and 1:1000 for anti-S or anti-E IgM and IgA. Horseradish peroxidase (HRP)-conjugated anti-human secondary antibodies were used at a 1:5000 dilution in 2.5% BSA in PBST: IgG1, IgG2, IgG3, and IgG4 (SouthernBiotech, Birmingham, AL, USA), IgM (Sigma-Aldrich), IgA (Invitrogen), and IgG (IgG H+L; Sigma-Aldrich). To establish standards for measuring antibody isotype concentrations, the following recombinant proteins were immobilized in carbonate buffer (0.1 M NaHCO3, pH 9.6) and serially diluted: IgM from serum (1:20,000, 2 mg/mL, Sigma-Aldrich), IgA from plasma (1:20,000, 1.44 mg/mL, Sigma-Aldrich), IgG1 heavy chain (1:1000, 0.25 mg/mL, Sino Biological), IgG2 heavy chain (1:5000, 0.25 mg/mL, Sino Biological), IgG3 heavy chain (1:20,000, 0.25 mg/mL, Sino Biological), and IgG4 heavy chain (1:10,000, 0.25 mg/mL, Sino Biological). For heavy chain standards, concentrations were adjusted to account for the lower molecular mass and the presence of two heavy chains in a complete IgG antibody.

### Anti-E Fc-receptor affinity assay

The envelope protein of Dengue virus type 2 (Sino Biological, Beijing, China) was immobilized at 0.2 mg/mL per well in a 96-well flat-bottom polystyrene microplate using bicarbonate buffer (NaHCO3, pH 9.6) and incubated overnight. A blocking solution of 2.5% bovine serum albumin (BSA) in TBST (TBS with 0.05% Tween 20) was applied to reduce nonspecific binding. After washing the plates with TBST, samples diluted 1:100 in TBST were added and incubated for 1 hour. The plates were washed four times with TBST, followed by the addition of biotinylated Fc-receptor CD16a or CD32a (AcroBiosystems) and streptavidin-alkaline phosphatase (Promega, diluted 1:10,000) in 2.5% BSA blocking solution for 1 hour. The plates were then washed nine times, and 50 µL of highly sensitive Tn-Cyclon reagent (BioPhenoMA) was added for signal detection. Absorbance at 405 nm was measured kinetically over 1 hour. The maximum reaction rate was used to quantify the signal, and the fold change was calculated relative to nonspecific IgG from human serum.

### Removal of anti-spike or anti-E antibodies

BcMag^TM^ Tosyl-Activated Magnetic Beads (BioClone, San Diego, CA, USA) were utilized to deplete anti-spike (anti-S) or anti-envelope (anti-E) antibodies. For antigen coupling, 100 μg of full spike or envelope protein (Sino Biological, Beijing, China) was conjugated to 30 mg of Tosyl magnetic beads. The beads were then evenly distributed across samples, with each sample receiving beads coupled with 2.5 μg of antigen. Serum or plasma samples were diluted at least 1:2000 in PBS, and 600 μL of diluted sample was mixed with 10–20 μL of beads, followed by incubation with over-the-top rotation at room temperature for 2 hours. The supernatant was collected and analyzed via ELISA or antibody-dependent enhancement (ADE) assays. Data were included only if the corresponding antibody titer was reduced by at least 80%. If the reduction was less than 80%, the experiment was repeated with higher sample dilutions (1:4000, 1:8000, or 1:10,000) until an 80% or greater reduction was achieved; otherwise, the data were excluded. The specificity of this method was confirmed using BSA-coated beads as a control, which resulted in a non-significant reduction in anti-S or anti-E antibody reactivity (9%, p=0.06 and 24%, p=0.06, respectively, **Supplemental figure 3 H, I**).

### ADE assay for serum antibodies

Dengue virus type 2 was incubated at a multiplicity of infection (MOI) of 0.001 with serum diluted 1:1000 for 30 minutes. The mixture was then incubated with 10^5 THP-1 cells for 90 minutes. The THP-1 cells were washed twice, and the supernatant was collected after 48 hours for analysis by quantitative PCR (qPCR).

#### qPCR of Dengue 2 virus

Reverse transcription was performed using 5 μL of total RNA in 20 μL reactions with the Reverse Transcription Mix (RT Enzyme & Kit, PURIGO Biotechnology Co., Ltd.) following the manufacturer’s instructions. Quantitative PCR (qPCR) was conducted on a LightCycler® 480 Instrument II (Roche) in 10 μL reactions using TaqMan™ Fast Advanced Master Mix (Applied Biosystems™, catalog #4444556), ∼1 μL of sample cDNA, and 10 μM each of forward and reverse primers (Dengue primers from Taiwan CDC). The qPCR protocol included pre-incubation at 95°C for 3 minutes, followed by 45 cycles of amplification (denaturation at 95°C for 10 seconds, annealing at 60°C for 20 seconds, and extension at 72°C for 5 seconds). After a final extension, a melting curve analysis was performed (95°C for 5 seconds, 60°C for 20 seconds, and cooling at 40°C for 30 seconds). All samples were run in technical duplicates, and expression levels were quantified as cycle quantification (Cq) values. For qPCR analysis Nike Air Max 97, the ΔΔCt method was applied, with gene expression fold changes expressed as log₂ values.

### Complement inactivation

To inactivate complement, the serum was diluted 1:500 in RPMI medium and incubated at 56°C for 30 minutes. For the complement rescue experiment, guinea pig complement (MP Biomedicals, Santa Ana, CA, USA; catalog #086428-CF) was added at a 1:50 dilution following heat treatment. Previous studies have shown that guinea pig complement can interact with and be activated by human antibodies with similar efficiency^11^.

### Statistics and data analysis

Statistical analysis was performed with R software (ver. 4.3.1) using the dplyr package. All figures were also created with R. For violin plots used in the presentation the median and interquartile difference is given. The Mann–Whitney U test was used for comparison between two samples. The pairwise Wilcoxon test was used to compare multiple groups. P values < 0.05 were considered significant. The pearson correlation was measured with the cor.test function in R was used to compare the significance of correlations of the IgG profile parameter with other variables (age, Fc-affinity, ADE and time interval from injection). To classify the cross-reactive antibody response in Figure 3, the mean and standard deviation of the differences between duplicate samples from the anti-envelope (anti-E) ELISA were calculated. The threshold for classification was defined as the mean plus the standard deviation (mean + standard deviation = threshold). If |*untreated(sample x) - anti_S_removed(sample x)*| < *threshold* = no cross-reaction. If there is a significant cross-reaction, full cross-reaction is determined by *anti_S_removed(sample x) - anti_E_removed(sample x)* < *threshold*, otherwise it will classified as partial cross-reaction. The classification was validated by applying a k-means clustering algorithm (kmeans function in R) to the data, using three clusters to determine if they correspond to the three groups: no cross-reactivity, partial cross-reactivity, and full cross-reactivity.

## Results

### Anti-Dengue envelope antibodies are induced by COVID-19 vaccination with a distinct antibody isotype profile than Dengue recovered

We gathered 271 serum and plasma samples from a previous Taiwanese population-based serum to assess the titers of anti-SARS-CoV-2 spike (anti-S) and anti-envelope (anti-E) IgM antibodies. The samples were categorized by vaccination status: unvaccinated (0: n=14), vaccinated with one to three doses of COVID-19 vaccines targeting the Wuhan variant (1: n=44; 2: n=60; 3: n=90), including adenovector, mRNA, and protein subunit vaccine types (**Table 1**). Additionally, we collected samples from individuals who received an Omicron-targeted booster, resulting in a total of 4–7 COVID-19 vaccine doses (XBB and BA.4.5, omi, n=21, **Supplemental table 1**). Furthermore, we collected samples from 24 individuals in southern Taiwan who had recovered from the 2023 dengue outbreak (DENV; n=24), with samples obtained at a median of 14 days post-diagnosis (range: 5–21 days). The infections were identified as either dengue virus serotype 1 or serotype 2 (**Supplemental table 2**). Both anti-S and anti-E IgM antibody levels were significantly elevated in individuals vaccinated against COVID-19 after the first dose. Additionally, anti-E IgM levels showed a significant increase following the second dose (**Figure A1, B1**). The IgM titers for anti-S and anti-E were comparable, with the median anti-E titer reaching 94% of the median anti-S titer (median anti-S: 4.3 µg/ml; anti-E: 3.5 µg/ml). In individuals recovered from the 2023 dengue outbreak, neither anti-S nor anti-E IgM titers were significantly elevated. For IgA antibodies, both anti-S and anti-E titers increased with each additional vaccine dose and were significantly higher compared to the unvaccinated cohort (**Figure A2, B2**). In COVID-19 vaccinated individuals, the anti-E IgA antibody titer was at median 40% anti-S IgA titer (median anti-S: 5.9 µg/ml; median anti-E: 2.5 µg/ml). In dengue recovered individuals, the median anti-E IgA titer (5.9 µg/ml) was higher than that in COVID-19 vaccinated individuals. For IgG antibodies, anti-E IgG titers showed a slight but significant increase from the first to the second COVID-19 vaccine dose but remained a minor fraction (median 2%) compared to anti-S IgG titers (median anti-S: 182.6 µg/ml; median anti-E: 2.5 µg/ml), which increased with each subsequent dose (**Figure A3, B3**). In dengue recovered individuals, anti-E IgG antibody titers were significantly elevated, as expected (median: 22.1 µg/ml). In COVID-19 vaccinated individuals, both anti-S and anti-E antibody titers were elevated across IgM, IgA, and IgG isotypes. The anti-E antibody titers in COVID-19 vaccinated individuals were lower than those elicited by the dengue infection (**Figure A4, B4**). For the IgG subclasses specific to anti-S, titers of all subclasses (IgG1, IgG2, IgG3, and IgG4) were significantly higher in COVID-19 vaccinated individuals compared to the unvaccinated population. Additionally, anti-S IgG1, IgG2, and IgG4 titers increased with each subsequent vaccine dose, consistent with our previous findings (**Figure 1 C**)^12^. For anti-E IgG in the COVID-19 vaccinated there was only a significant increase of anti-E IgG1 in individuals who received omicron variant booster (**Figure 1 D1**), in IgG2 from the 1^st^ to the 2^nd^ dose and in IgG4 the was a significant increase for the 1^st^ and 3^rd^ dose (**Figure 1 D2,D4**). Dengue-recovered had a typical anti-viral anti-E IgG profile with significantly increased IgG1 and IgG3 (**Figure 1 D1, D3**). Overall, our findings indicate that the anti-E antibody profile in COVID-19 vaccinated individuals is dominated by IgM and IgA isotypes, with titers increasing with each vaccine dose. In contrast, the anti-E antibody profile in dengue recovered and the anti-S antibody profile in vaccinated individuals are predominantly IgG1-driven (**Figure 1 E-H**). No significant differences were observed in the anti-E antibody titers across IgM, IgA, and IgG isotypes among the different COVID-19 vaccine platforms (adenovector, mRNA, and protein subunit) in vaccinated individuals (**Supplemental figure 1**). For anti-SARS-CoV-2 spike (anti-S) antibody titers, significantly lower IgA and IgG subclass titers were observed in individuals vaccinated with the AstraZeneca (adenovector) vaccine compared to those vaccinated with mRNA vaccines, consistent with our previous findings ^12^. No significant correlation was observed between age and anti-S or anti-E antibody titers across IgM, IgA, and IgG isotypes, with the exception of anti-E IgG1, which showed a negative correlation with age following the first COVID-19 vaccine dose (**Supplemental Figure 2 A**). The interval between COVID-19 vaccination and sample collection was positively correlated with anti-E IgG2 titers following the first vaccine dose. Conversely, after the Omicron-targeted booster, the interval between vaccination and sample collection was negatively correlated with both anti-E IgM and anti-E IgG2 titers (**Supplemental Figure 2 B**). The sex had no significant influence on the antibody isotype titer (**Supplemental Figure 2 C-H**). An infection with SARS-CoV-2 increased the anti-S IgA, IgG1,2,4 titer in COVID-19 vaccinated, but had no effect on the anti-E antibody titer (**Supplemental Figure 2 C-H**).

**Figure 1:**
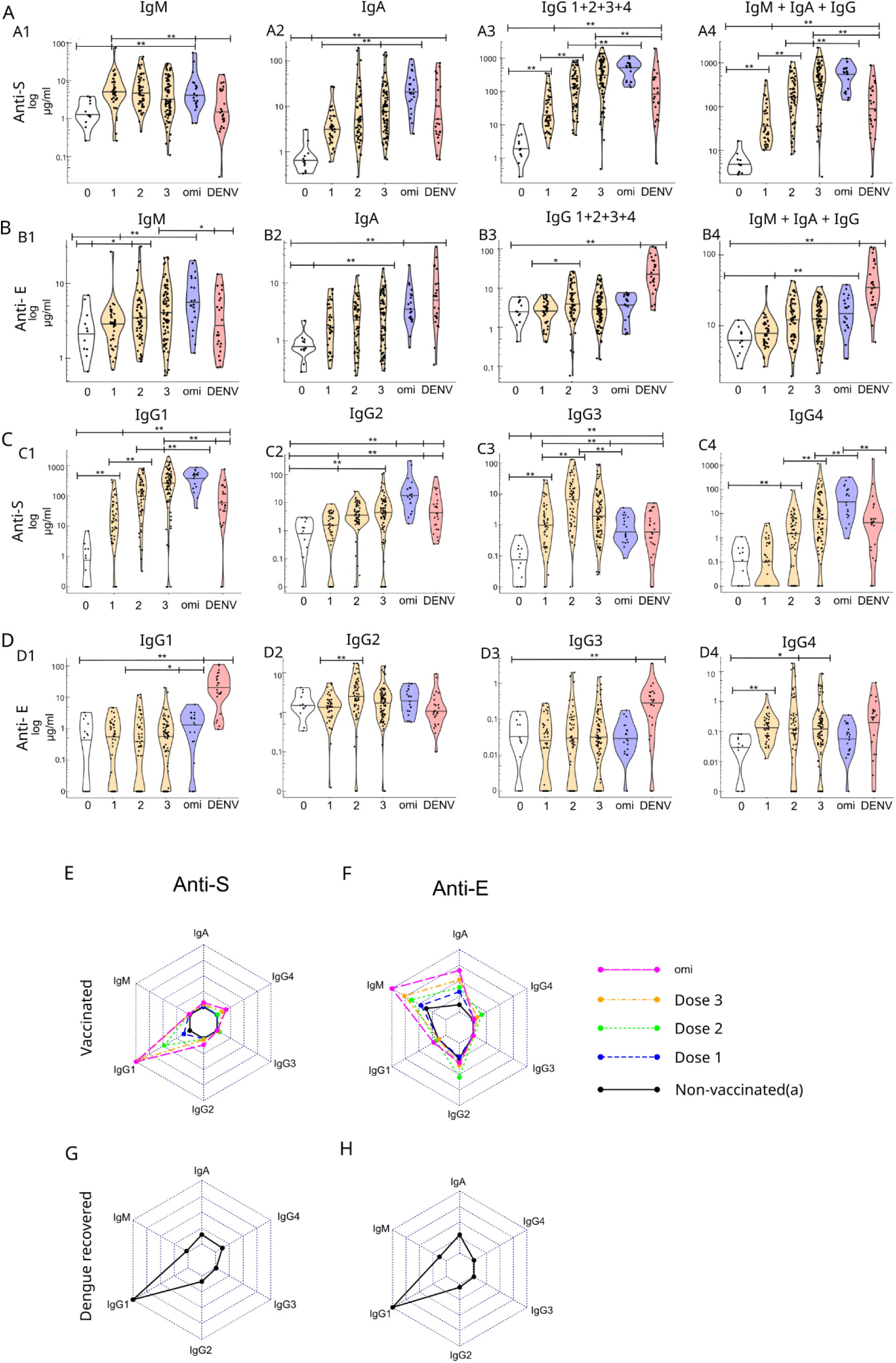
Antibody titers for anti-SARS-CoV-2 spike (anti-S) and anti-Dengue type 2 envelope (anti-E) were measured in unvaccinated, COVID-19 vaccinated, and dengue-recovered individuals. Titers are reported in µg/ml for each isotype (IgM, IgA, total IgG [IgG1+2+3+4], and the sum of all isotypes) in sections (**A-B**). Titers for anti-S and anti-E are further detailed for each IgG subclass (IgG1-4) in sections (**C-D**). The average contribution of each isotype to anti-S and anti-E responses in the COVID-19 vaccinated group is visualized in radar plots (**E-F**), and for the dengue-recovered group in radar plots (**G-H**). Radar plots are normalized to the highest average isotype concentration. Statistical significance is denoted as *p<0.01.

### Low anti-E and anti-spike titer are present before vaccination and are significantly differently between ethnicities/regions

We detected low levels of anti-S and anti-E antibodies in the unvaccinated Taiwanese cohort (**Figure 2**). Given that flaviviruses, such as dengue and Japanese encephalitis, are endemic in Taiwan and the samples were collected after the onset of the COVID-19 pandemic, we investigated whether these low antibody levels were specific to Taiwan by comparing anti-S and anti-E antibody titers with pre-pandemic samples from the United States. The U.S. samples also exhibited detectable anti-S and anti-E titers. No significant differences were found in anti-S and anti-E IgM titers between the two cohorts (**Figure 2A**). The U.S. cohort showed significantly higher anti-S and anti-E IgA titers (**Figure 2B**), while the Taiwanese cohort had higher anti-S/E IgG titers, with significantly elevated levels for IgG2, IgG3, and IgG4 subclasses (**Figure 2C–F**).

**Figure 2:**
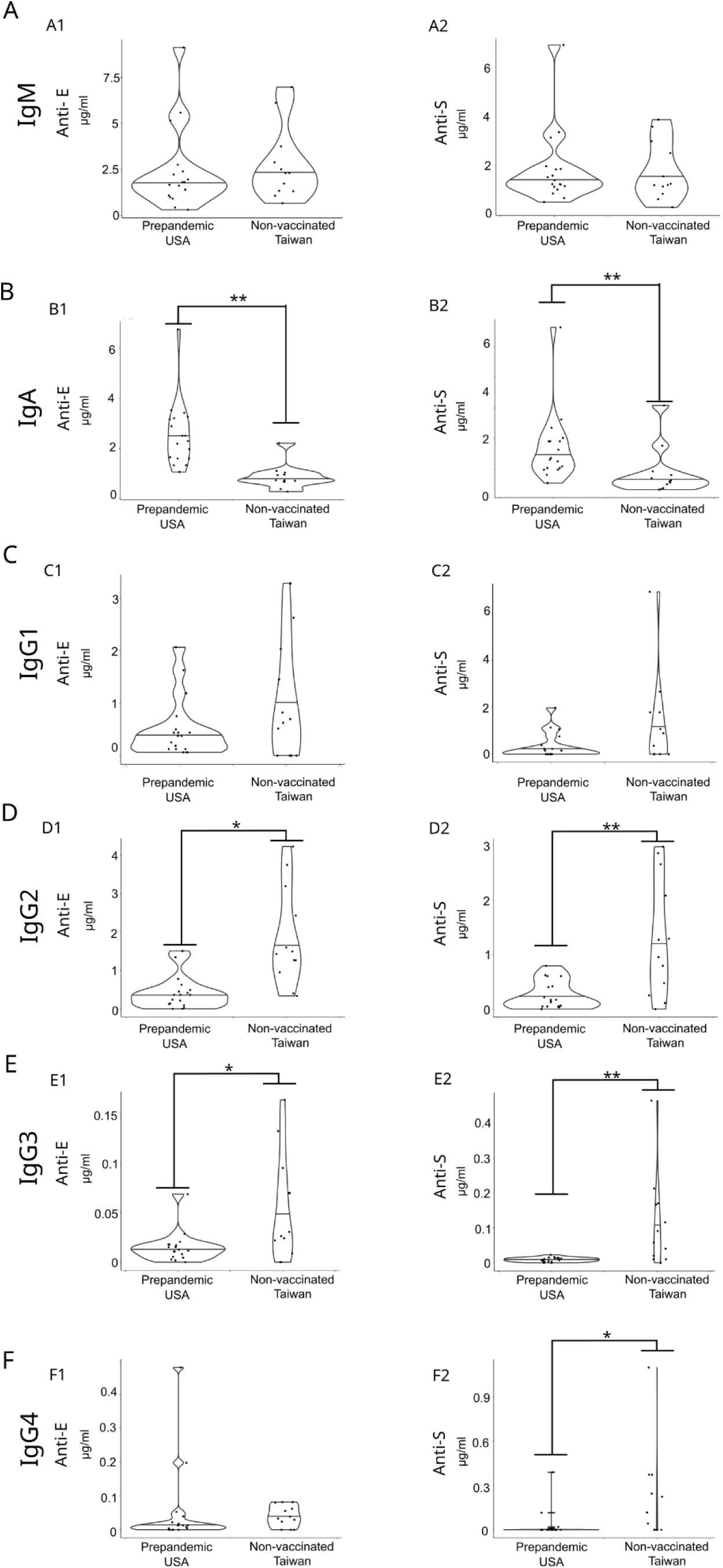
Concentrations of anti-E and anti-S antibodies were measured in non-vaccinated Taiwanese and pre-pandemic U.S. cohorts for the following isotypes: (**A**) IgM, (**B**) IgA, and (**C-F**) IgG subclasses. Statistical significance is indicated as *p<0.05 and **p<0.01., * p<0.05, ** p<0.01

### Anti-E antibodies from COVID-19 vaccinated are fully cross-reactive to the SARS-CoV-2 spike protein

In individuals vaccinated against COVID-19, we observed significant correlations between anti-S and anti-E antibody titers for both IgM and IgA isotypes (**Figure 3A**). In contrast, among individuals recovered from dengue infection, only IgM titers showed a significant correlation (**Figure 3B**). In the unvaccinated Taiwanese cohort, cross-correlations between anti-S and anti-E antibody titers were stronger than in the vaccinated cohort. Significant correlations were observed for IgM, IgA, and IgG1 isotypes. Additionally, a significant correlation was found between anti-S IgG1 and anti-E IgG2 titers (**Supplemental Figure 3 A**). To investigate whether elevated anti-E antibodies in COVID-19-vaccinated individuals cross-react with the SARS-CoV-2 spike protein, we selectively depleted either anti-S or anti-E antibodies using magnetic beads coated with S or E proteins, respectively. If anti-E antibodies are cross-reactive with anti-S antibodies, depletion of anti-S antibodies would also reduce anti-E titers. Our results confirmed cross-reactivity, as depletion of anti-S antibodies led to a reduction in anti-E titers for IgM, IgA, and IgG isotypes. (**Figure 3 C-E**). In all COVID-19-vaccinated individuals (third dose, n=19) and dengue-recovered individuals (n=12), we confirmed cross-reactivity for IgM antibodies in all investigated samples. The degree of cross-reactivity was quantified by calculating the relative difference between anti-S and anti-E antibody titers. We observed a high level of cross-reactivity, with no significant difference in IgM cross-reactivity between the COVID-19-vaccinated and dengue-recovered groups (**Figure 3 C4**). Furthermore, depletion of anti-S antibodies significantly reduced anti-S titers to a comparable extent across all samples (**Supplemental Figure 3 B, C**), indicating that most anti-S and anti-E antibodies are cross-reactive. In contrast, depletion of anti-E antibodies resulted in only a minor or non-significant reduction in anti-S titers for IgA and IgG isotypes (**Supplemental figure 3 D-G**), indicating that most of the anti-S are not cross-reactive to anti-E either in COVID-19 vaccinated or Dengue recovered. Anti-E IgA and IgG antibodies exhibited a higher degree of cross-reactivity in COVID-19-vaccinated individuals compared to dengue-recovered individuals (**Figure 3 D**). Notably, one individual in the vaccinated cohort and two individuals in the dengue-recovered cohort showed no detectable cross-reactive anti-E IgG antibodies and one dengue recovered did not show cross-reactivity for IgA (**Figure 3 D, E**).

**Figure 3:**
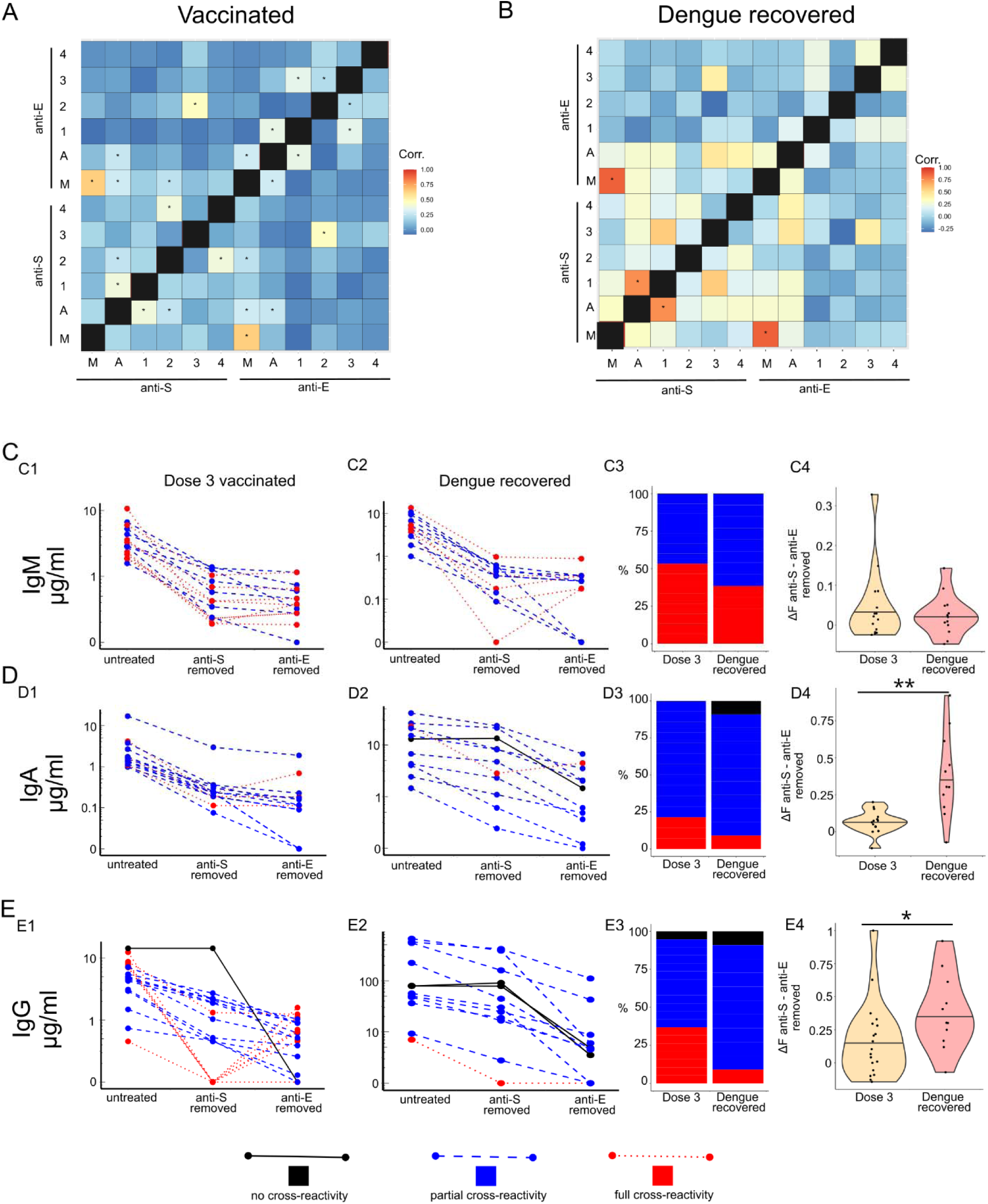
(**A-B**) Pearson correlation between anti-spike an anti-E Isotypes in COVID-19 vaccinated individuals. (**C-E**) Anti-E titer before (untreated) or after removal of anti-S or anti-E antibodies with magnetic beads either from COVID-19 vaccinated (3 dose) or Dengue recovered individuals. **(C1-E2)** the direct titer of vaccinated and Dengue recovered after anti-S and anti-E removal. Samples classified as non-cross reactive are marked as black with solid lines, partial cross-reactive blue with dashed lines and full cross-reactive as red with dotted lines. **(C3-E3)** Distribution of non-, partial and fully cross-reactive anti-E samples. **(C4-E4)** Level of cross-reactivity given as 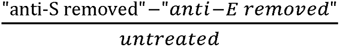. C * p<0.05, ** p<0.01

### Antibodies from COVID-19 vaccinated can enhance the infection of Dengue virus

In the next step we want to test the capability of the cross-reactive antibodies to induce antibody dependent enhancement (ADE) in dengue infections. We therefore incubate dengue viruses with serum/plasma samples and then test the enhancement of infection by Dengue virus in human monocyte like THP-1. There was a significant increase of ADE for the first dose and decreased then to the 3^rd^ dose (**Figure 4 A**). In individuals who received the Omicron booster, a significant ADE effect was observed. Similarly, antibodies from dengue-recovered individuals also exhibited a significant ADE effect. Another critical factor influencing the risk of severe secondary disease is the affinity of antibodies for the FcγRIIIa receptor^13^. Compared to dengue-recovered individuals, COVID-19-vaccinated individuals displayed lower binding affinity of anti-E antibodies to both FcγRIIa and FcγRIIIa receptors (**Figure 4 B,C**). No differences in FcγRIIIa binding affinity of anti-E antibodies were observed among the vaccines, except for the BNT162b2 vaccine, which exhibited significantly higher FcγRIIIa affinity after the first dose (**Supplemental Figure 3 B**). No significant correlation was observed between ADE and either antibody isotype or anti-E Fc-receptor affinity in COVID-19-vaccinated or dengue-recovered individuals, except for a significant correlation between FcγRIIa and FcγRIIIa affinities (**Figure 4 D,E**). Depletion of either anti-S or anti-E antibodies significantly reduced ADE in COVID-19-vaccinated individuals (**Figure 4 F**). In dengue-recovered individuals, depletion of anti-E antibodies resulted in a greater reduction of ADE compared to depletion of anti-S antibodies (**Figure 4 G**). Studies have shown that the complement system can suppress ADE ^9,14^. IgG1 and IgG3 exhibit stronger binding affinity for C1q, enhancing activation of the classical complement pathway^15^ and therefore sera from individuals recovered from dengue, with high IgG1 and IgG3 anti-envelope (anti-E) antibody titers, may exhibit stronger complement-mediated inhibition of ADE compared to sera from COVID-19-vaccinated individuals with elevated anti-E IgG2 and IgG4 titers. We inactivated the complement system by heat treatment at 56°C for 30 minutes. Complement inactivation significantly increased ADE activity in sera from dengue-recovered individuals, whereas it significantly reduced ADE activity in sera from individuals vaccinated with a single dose of a COVID-19 vaccine exhibiting high ADE activity. (**Figure 4 H,I**). Additionally, supplementation with active complement restored the reduced antibody-dependent enhancement (ADE) activity in sera from dengue-recovered individuals but had no effect on ADE activity in sera from COVID-19-vaccinated individuals (**Figure 4 H,I**).

**Figure 4:**
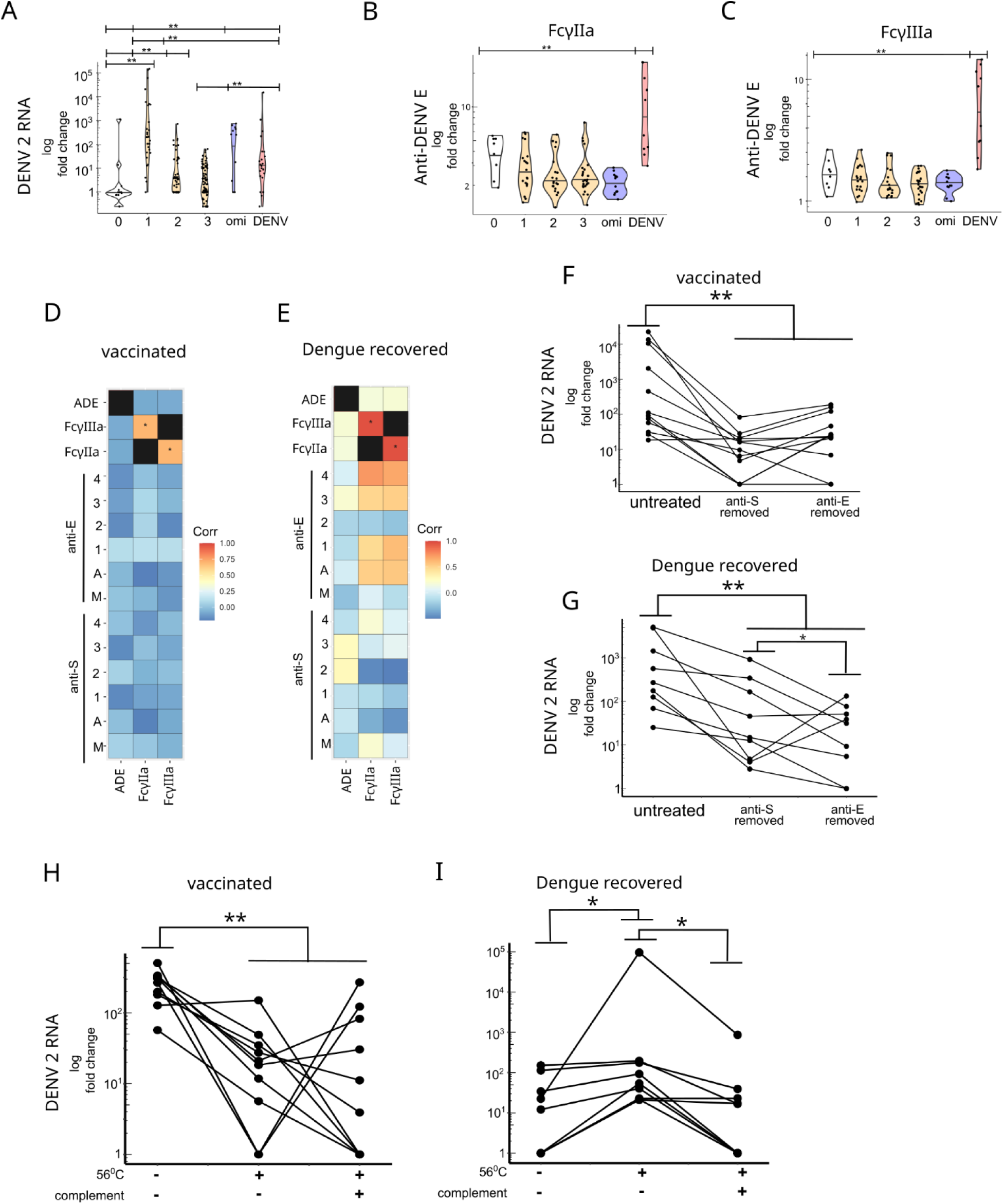
(**A**) Antibody dependent enhancement in THP-1 monocytes. Viral replication is measured by qPCR for Dengue RNA. Fold change is the difference to virus only treatment.(**B-C**) Affinity of anti-E antibodies to FcγIIa and FcγIIIa IgG receptor. Fold change is measured as ratio to the signal for IgG isotype unspecific for Dengue. (**D-E**) correlation of ADE, FcγIIa and FcγIIIa to anti-spike or anti-E antibody isotypes. (**F-G**) ADE measured by qPCR before and after removal of anti-spike or anti-E antibodies. **(H-I)** ADE measured in untreated versus complement inactivated sera (56^0^C) and complement inactivated versus guinea pig complement added samples in COVID-19 vaccinated (1 dose) and COVID-19 recovered individuals. * p<0.05, ** p<0.01

## Discussion

Our research provides the first characterization of distinct antibody isotype profiles for cross-reactive antibodies between SARS-CoV-2 and dengue virus. Although previous studies have reported cross-reactivity of antibodies from infected or vaccinated individuals between these viruses, no prior work has specifically examined differences in antibody isotype distribution in response to SARS-CoV-2 antigens or in COVID-19-vaccinated individuals. Additionally, few studies have explored epitope specificity for cross-reactive antibodies between SARS-CoV-2 and dengue. One study identified 24 potential IgA epitopes on the SARS-CoV-2 spike protein using in silico sequence comparison ^16^. However, the referenced study did not provide direct evidence for the presence of cross-reactive antibodies and failed to detect cross-reactivity for some proposed peptide epitopes in dengue-recovered patients. Several studies have explored the isotype profiles of cross-reactive antibodies between dengue and other flaviviruses. For instance, one study examining antibody responses in dengue patients to Japanese encephalitis virus (JEV) and dengue virus non-structural protein 1 (NS1) found no significant differences in the distribution of IgM, IgA, and IgG isotypes between the two viruses ^17^. study of Zika-infected individuals identified a significant role for IgM cross-reactive antibodies in virus neutralization, with one individual also exhibiting a notable IgA contribution to neutralization^18^. Another study focusing solely on dengue virus reported that each IgG subclass targets distinct domains of the dengue envelope (E) protein following prolonged seroconvalescence^19^.

Cross-reactive antibodies are present in unvaccinated cohorts from both the United States and Taiwan. One possible explanation is that these antibodies originate from natural antibodies expressed without prior antigenic stimulation, although their levels may increase following antigenic exposure^20^. Natural antibodies are predominantly of the IgM class, and most anti-E and anti-S IgM antibodies exhibit cross-reactivity across all cohorts. One study reported that dengue virus infection activates natural IgM, IgA, and IgG antibodies. This study also demonstrated that dengue virus infection induces antibodies against unrelated viruses, such as poliovirus^21^. Further investigation of B cells producing anti-E and anti-S antibodies is needed to test the hypothesis. Our study could not identify the reasons for differences in cross-reactive antibody profiles between unvaccinated Asian Taiwanese and predominantly Black US cohorts. Beyond genetic factors, environmental influences, such as prior antigenic exposure, may contribute, particularly since childhood Japanese encephalitis (JE) vaccination in Taiwan could account for the elevated anti-E IgG titers in the Taiwanese population^22^.

Our study did not investigate the epitopes of cross-reactive antibodies or whether epitope specificity varies across different antibody isotypes. Studies on HIV and a bacterial pathogen have shown that the constant domain of distinct IgG isotypes can affect the conformation of the Fab variable region, thereby influencing epitope specificity^23–25^. Studies on autoimmune diseases and allergies have shown that different antibody isotypes target distinct epitopes^26–28^. Peptide-based epitope mapping could identify cross-reactive epitopes, as previously demonstrated by the discovery of a shared epitope between the dengue virus envelope (E) protein and the SARS-CoV-2 spike protein receptor-binding domain (RBD)^4^. The identified cross-reactive epitope contains a glycosylation site, and the predominant cross-reactive antibodies in our study were IgM, with IgG2 as the primary IgG subclass. Both IgM and IgG2 can be produced through T-cell-independent mechanisms and may target glycan epitopes. Future research should include not only peptide-based epitope mapping but also analysis of glycopeptide targets to further elucidate cross-reactivity.

Our study confirmed that cross-reactive antibodies induce vitro ADE, despite their low titers and reduced Fc receptor affinity compared to antibodies in dengue-recovered individuals. This may be partly explained by differences in complement system involvement. Complement components C1q and C3 can inhibit ADE, and, as expected, dengue-recovered individuals exhibited lower ADE due to higher levels of anti-E IgG1 and IgG3, which have greater affinity for C1q than IgG2 and IgG4, leading to ADE inhibition either directly via C1q or through C3 activation ^9,14^, which fragment can deposit on the surface of the Dengue virus and either can trigger the direct lysis or subject the virion to another phagocytosis pathway via C3R receptor. Unexpectedly, complement inactivation reduced ADE activity in SARS-CoV-2-vaccinated individuals, warranting further investigation into the role of the complement system in low-titer anti-dengue IgG2 and IgG4 antibodies or the involvement of other temperature-sensitive serum proteins. While 1^st^ dose and omicron targeted booster caused a larger in vitro ADE than dengue recovered, there is no study that saw an association with COVID-19 vaccination and dengue disease. The observed effects may be explained by low IgG1 titers and reduced FcγRIIIa affinity, as low-fucosylated IgG1 with high FcγRIIIa affinity has been associated with severe secondary dengue disease in observational and animal studies ^13,29,30^. Therefore, these cross-reactive low FcγRIIIa affinity antibodies from COVID-19 vaccines are no risk factor for more severe dengue disease.

Previous studies suggested that the omicron variant or omicron-specific booster may reduce titers of cross-reactive anti-dengue antibodies for dengue virus serotype 2^5^. However, our study found that the omicron-boosted cohort had the highest anti-E antibody titers among all vaccinated cohorts, accompanied by elevated ADE activity. Additionally, omicron-boosted individuals showed increased anti-E IgG1 levels. A negative correlation was observed between the time interval from omicron booster administration to sample collection and anti-E titers, suggesting a potential negative effect of the omicron booster on cross-reactive antibodies in the long term. Studies, which investigate the long-term impact of the omicron targeted booster on cross-reactive antibodies would be necessary.

### Conclusion

Our study demonstrated that cross-reactive antibodies against dengue virus, induced by SARS-CoV-2 vaccination, exhibit a distinctive IgM- and IgA-dominated isotype profile. Despite low anti-dengue IgG titers, these antibodies were capable of inducing antibody-dependent enhancement (ADE). The mechanisms underlying these effects remain unclear and warrant further investigation.

## Limitations

Our observational study lacked a fixed time interval between sample collection and vaccination, resulting in variable sampling times. No significant differences were observed in age or gender distribution, but data on comorbidities were not recorded. For omicron-booster-vaccinated individuals, prior vaccination histories were inconsistent, with some participants unable to provide complete COVID-19 vaccination records. Our dengue-recovered cohort, collected in 2023, reflects a Taiwanese population where most individuals were either vaccinated against or exposed to SARS-CoV-2, precluding data on cross-reactive antibodies in dengue-recovered individuals unexposed to SARS-CoV-2. In the antibody removal assay for anti-spike (S) and anti-envelope (E) antibodies, samples with very low anti-E or anti-S titers were excluded due to the high dilution required, while high-titer samples risked incomplete antibody removal, potentially introducing bias toward samples with specific titer ranges.

## Data Availability

Data can be provided upon request.

## Acknowledgements

We thank Dr.Hsing I-Huang and Chi-Chong Chio for providing the THP-1 cell line. This work has been supported by NIH NIAID U01AI151698, United World Antiviral Research Network, part of the NIAID CREID network. This work was also supported in part by the Research Center for Emerging Viral Infections from The Featured Areas Research Center Program within the framework of the Higher Education Sprout Project by the Ministry of Education (MOE) in Taiwan. Additionally, we received support from the Ministry of Science and Technology (MOST), Taiwan (grant number: MOST 111-2634-F-182-001, MOST 109-2327-B-182-002), and the Chang Gung Memorial Hospital (grant number: CORPD1K0011-12, CLRPD1J0015).

## Conflict of interests

Etsuro Ito is member of the R&D Department, BioPhenoMA Inc., Tokyo, Japan.

## Contribution

Sebastian Reinig developed and performed the antibody isotype ELISA. The anti-S/E depletion assay was developed and performed by Sebastian Reinig and Kou-Chin. The Fc-affinity assays were developed by Po-kai Chen, Etsuro Ito, Sebastian Reinig and Kou-Chin and finally performed by Sebastian Reinig. The qPCR was performed by Kou-Chin. Antibody enhancement assays and cell culture maintenance were performed by Kou-Chin and Sebastian Reinig. Sheng-Yu Huang and Kuei-Ching Hsiung collected the serum/plasma samples and determined the dengue serotype by neutralization assay. Ing-Kit Lee, Ching-Yen Tsai, Shu-Min Lin assisted in recruiting eligible participants. Sebastian Reinig created the figures and performed the data analysis. The manuscript was written and edited by Sebastian Reinig, Kuei-Ching Hsiung, Sheng-Yu Huang and Shin-Ru Shih.

## Ethics statement

This study was conducted under the approval and oversight of the Institutional Review Board (IRB) of the Chang Gung Medical Foundation, based in Linkou, Taiwan (IRB approval number: 202001041A3C). The IRB is responsible for the ethical review of all human subject research conducted at Chang Gung Memorial Hospital, Chang Gung University, and Chang Gung University of Science and Technology. The IRB follows the guidelines of the Taiwan Ministry of Health and Welfare and operates in accordance with the Declaration of Helsinki. In addition, sample collection and participant enrollment at Linkou Chang Gung Memorial Hospital, Kaohsiung Chang Gung Memorial Hospital, and Kaohsiung Municipal Fengshan Hospital were reviewed and approved by the same IRB under the same protocol. All participants provided written informed consent in accordance with local legal and ethical standards, including those outlined in Taiwan’s Human Subjects Research Act and Medical Care Act.The study is part of the University of Washington Arboviral Research Network (UWARN), supported by the U.S. National Institutes of Health (project number: 1 U01 AI151698). The Principal Investigator is Prof. Shin-Ru Shih (Chang Gung University, Research Center for Emerging Viral Infections). The site Co-Principal Investigators in southern Taiwan are Dr. Ing-Kit Lee (Kaohsiung Chang Gung Memorial Hospital) and Dr. Ching-Yen Tsai (Department of Infectious Diseases, Kaohsiung Municipal Fengshan Hospital). The site Co-Principal Investigator in northern Taiwan is Dr. Shu-Min Lin (Linkou Chang Gung Memorial Hospital).

