## Supplemental Material for "COVID-19 Vaccination Induces Cross-Reactive Dengue Antibodies with Altered Isotype Profiles and In Vitro ADE"

**Supplemental table 1**: Booster vaccines and prior vaccination history of the samples of the omicron targeted cohort. ChAdOx1 nCoV-19 (AstraZeneca, AZ), MVC-COV1901 (Medigen), or the mRNA vaccines mRNA-1273 (Moderna), BNT162b2 (BNT). Peptide: Experimental peptide vaccine from United Biomed, Protein subunit Novavax: NVX-CoV2373

| Omicron booster | dose | Prior vaccinated |
| --- | --- | --- |
| Novavax (XBB) | 5 | AZ-AZ-BNT-unknown |
|  | 5 | AZ-AZ-Moderna-unknown |
|  | 5 | Moderna-Moderna-unknown-unknown |
|  | 4 | Unknown |
|  | 4 | AZ-AZ-BNT |
|  | 7 | United Biomed-United Biomed-Medigen-Medigen-BNT-BNT |
|  | 5 | Moderna-Moderna-Medigen-Novavax |
|  | 7 | Medigen-Medigen-Medigen-Medigen-Moderna-Moderna |
|  | 8 | Peptide-Peptide-unknown |
|  | 5 | AZ-AZ-BNT-unknown |
|  | 5 | AZ-AZ-Medigen-unknown |
|  | 5 | AZ-AZ-Moderna-Novavax |
|  | 5 | unknown |
| Moderna (XBB) | 5 | AZ-AZ-Moderna-unknown |
|  | 4 | BNT-BNT-BNT |
|  | 5 | AZ-AZ-Moderna-Moderna(BA.4.5) |
|  | 6 | Medigen-Medigen-BNT-Moderna(BA.1)-Moderna(BA.4.5) |
|  | 5 | AZ-AZ-Moderna-Moderna(BA.1) |
| Moderna (Ba.4.5.) | 4 | AZ-AZ-Moderna |
|  | 4 | BNT-BNT-BNT |

**Supplemental table 2**: Details of the Dengue recovered cohort. ChAdOx1 nCoV-19 (AstraZeneca, AZ), MVC-COV1901 (Medigen), or the mRNA vaccines mRNA-1273 (Moderna), BNT162b2 (BNT).

| **ID** | **age** | **sex** | **Interval**  **(days from**  **diag-nosis)** | **serotype** | **Number of symptoms** | **Prior COVID-19**  **vaccination** | **COVID**  **infected** |
| --- | --- | --- | --- | --- | --- | --- | --- |
| 2 | 41-45 | F | 13 | 1 | 7 | ND | no |
| 3 | 31-35 | F | 18 | 1 | 7 | ND | yes |
| 4 | 61-65 | M | 15 | ND | 7 | ND | yes |
| 5 | 81-85 | F | 17 | 1 | 5 | ND | no |
| 6 | 76-80 | F | 20 | 1 | 8 (bleeding gum) | ND | no |
| 7 | 66-70 | M | 9 | 1 | 8 | Moderna-Moderna-Moderna | no |
| 10 | 36-40 | M | 10 | 1,2 | ND | ND | yes |
| 11 | 21-25 | F | 18 | 1 | ND | ND | yes |
| 12 | 61-65 | M | 12 | 1 | ND | AZ-AZ-Moderna-Moderna | no |
| 13 | 66-70 | M | 18 | ND | ND | Moderna-Moderna | yes |
| 14 | 66-70 | M | 18 | 1 | ND | ND | yes |
| 15 | 81-85 | F | 16 | 1 | 4 | ND | no |
| 16 | 21-25 | M | 14 | ND | ND | BNT-BNT-Moderna-Moderna | no |
| 17 | 41-45 | F | 12 | ND | ND | BNT-BNT | no |
| 18 | 51-55 | M | 10 | 2 | 4 | ND | no |
| 19 | 66-70 | M | 12 | ND | ND | AZ-AZ-Moderna-Moderna | no |
| 20 | 56-60 | F | 21 | 2 | ND | ND | yes |
| 21 | 56-60 | F | 12 | 1 | ND | ND | yes |
| 22 | 21-25 | M | 13 | ND | ND | Medigen | yes |
| 23 | 46-50 | F | 14 | ND | ND | ND | no |
| 28 | 51-55 | M | 5 | ND | ND | Moderna-Moderna-Moderna | yes |

**Supplemental table 3:** Details of the unvaccinated and Pre-pandemic US cohort.

|  | Pre-pandemic (USA) | Non-vaccinated (Taiwan) |
| --- | --- | --- |
| n | 18 | 14 |
| Collection period | 2017/4/28-2018/11/9 | 2021/8/19-2023-2022/6/7 |
| age | 39.05 (22-64) | 37.2 (21-59) |
| Female(%) | 66.6 | 71.4 |
| ethnicity | Black (16), Caucasian (1), Hispanic (1) | Asian (14) |

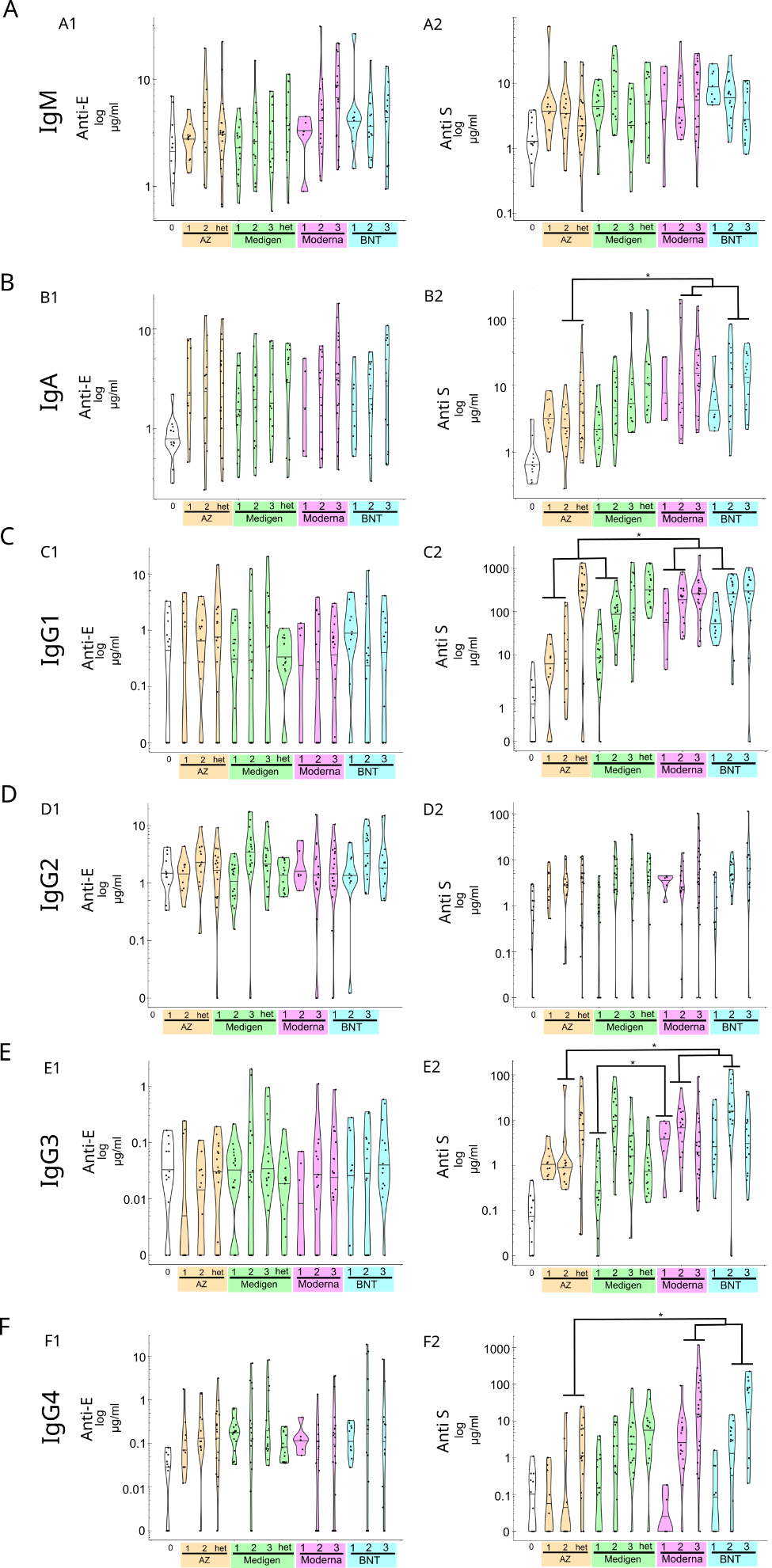

**Supplemental Figure 1:** anti-SARS-CoV-2 spike (anti-spike) antibody titer or anti-Dengue type 2 envelope (Anti-E) titer between individuals vaccinated with different vaccine platforms. The antibody titer is given in µg/ml for each Isotype **(A)** IgM, IgA **(B)** and **(C-F)** IgG1-4. * p<0.05, ** p<0.01, het: heterology (1-2^nd^ dose non-mRNA and 3^rd^ dose mRNA vaccine).

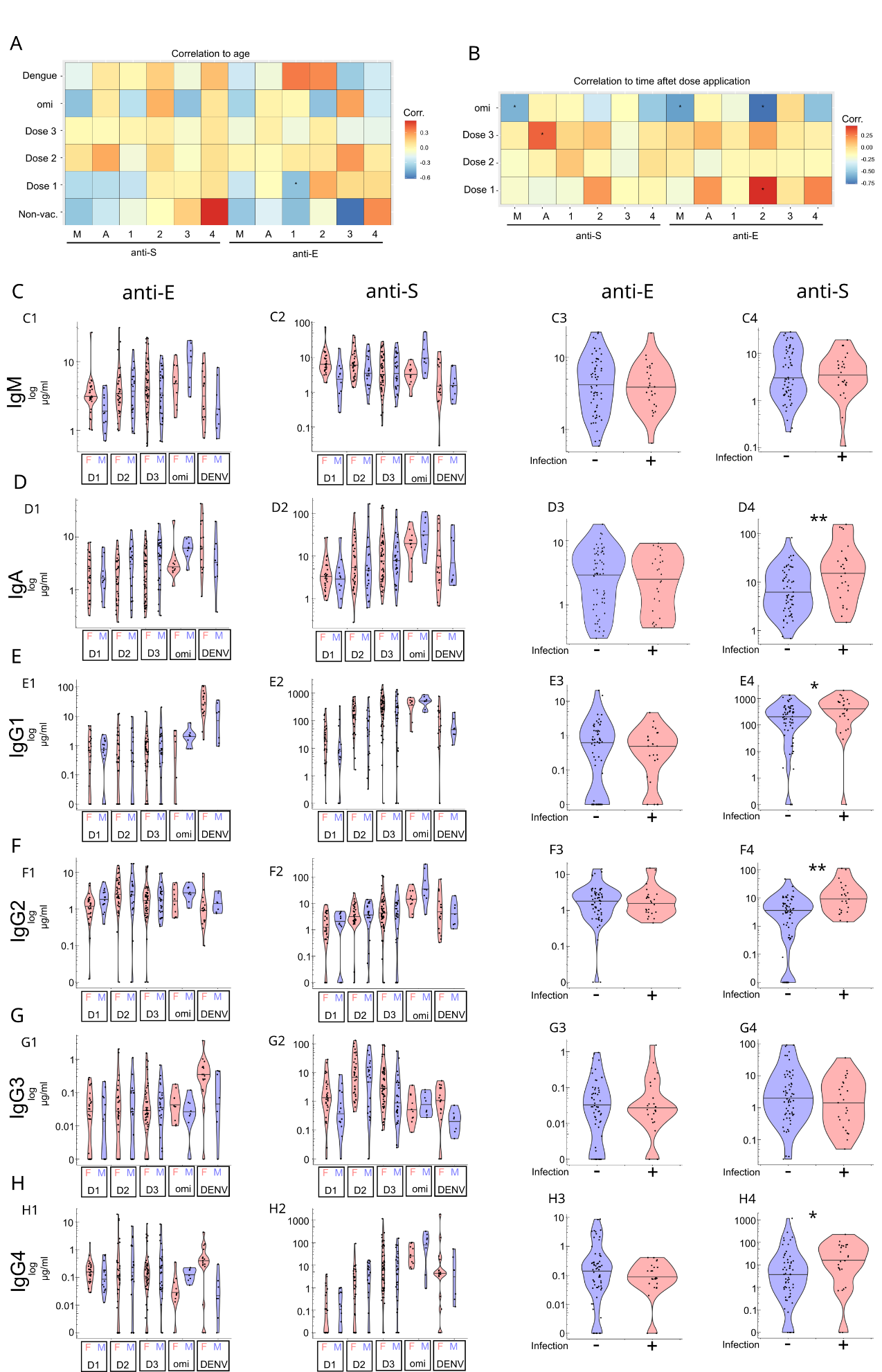

**Supplemental Figure 2: (A)** Correlation of antibody isotypes and cohorts to age or **(B)** the time interval after the last vaccine dose. (**C-H**) anti-S and anti-E Isotype titer separated by sex (F: Female and M: Male) or COVID-19 infection. For infection only individuals with the 3^rd^ dose are are included due to the lack of COVID-19 recovered individuals in the 1^st^ and 2^nd^ dose cohort. * p<0.05, ** p<0.01

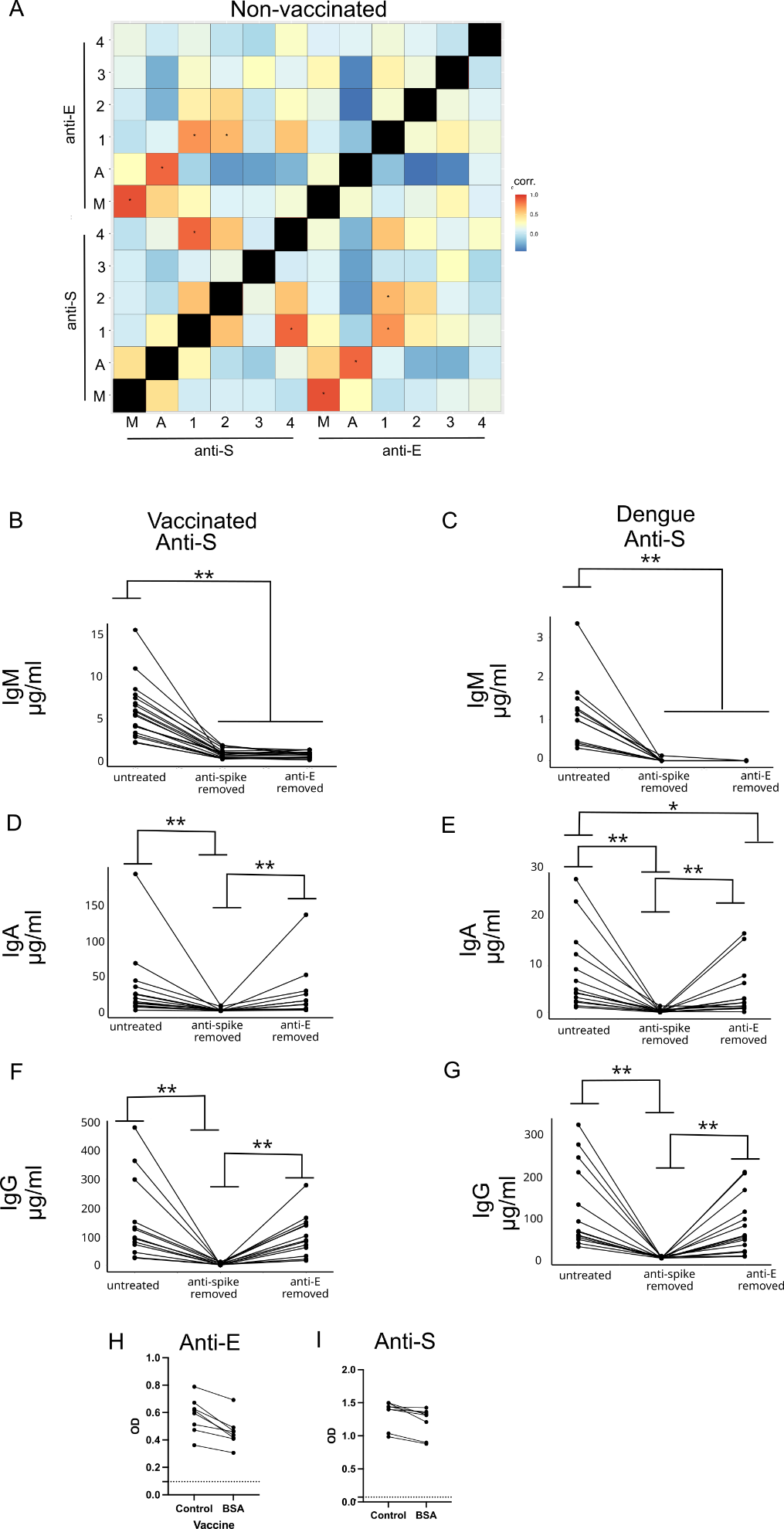

**Supplemental Figure 3:** **(A)** Pearson correlation between anti-S and anti-E of all unvaccinated individuals between different isotypes. (**B-G**) anti-S titer before and after removal of anti-S or anti-E antibodies with magnetic beads either from COVID-19 vaccinated (3 dose) or Dengue recovered individuals. **(H)** Anti-S and **(I)** Anti-E titer of beads before and after treatment with BSA coated beads to test specificity of antigen removal of antibodies. * p<0.05, ** p<0.01

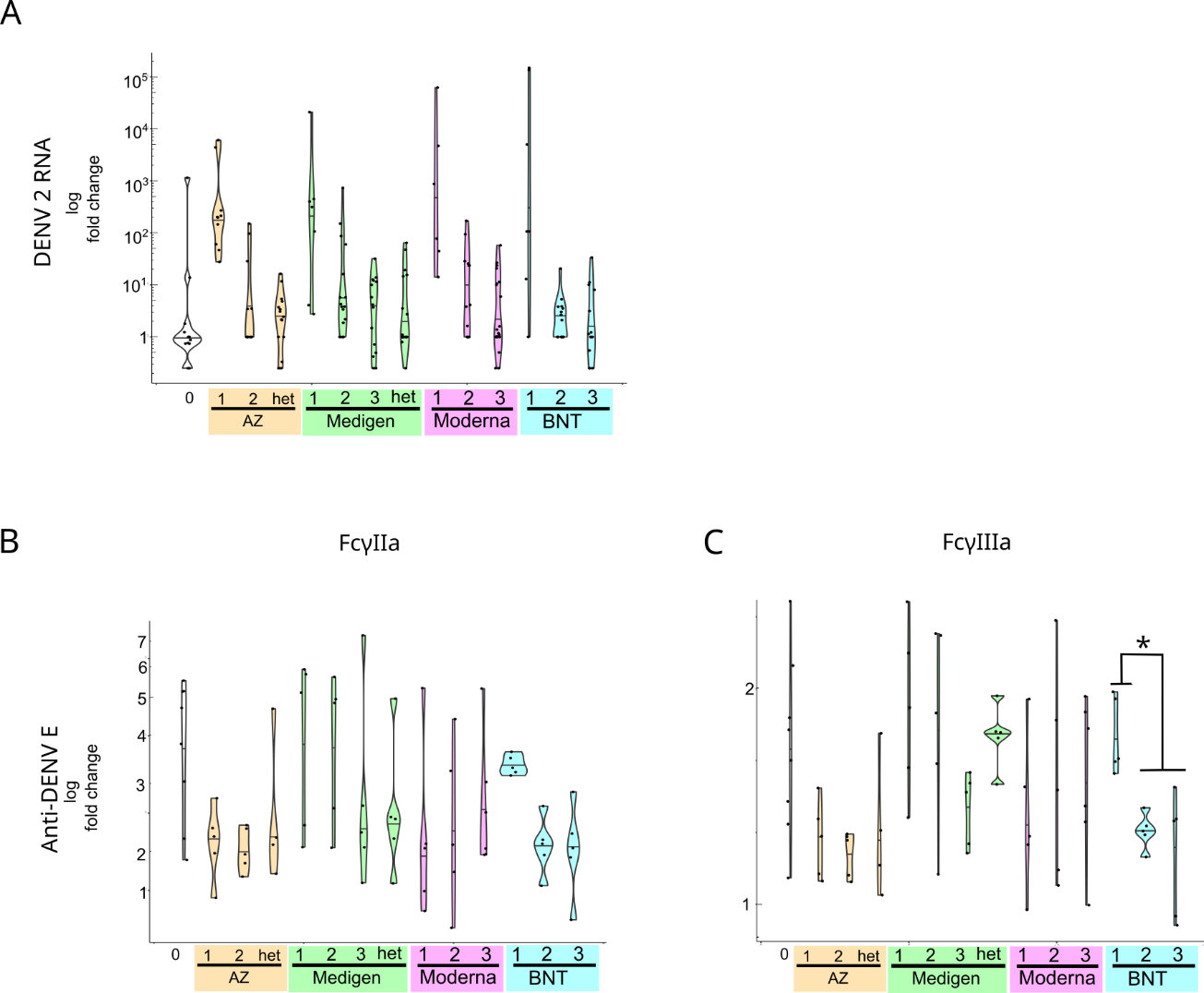

**Supplemental Figure 4**: **(A)** Antibody dependent enhancement in THP-1 monocytes for each vaccine platform. Viral replication is measured by qPCR for Dengue RNA. Fold change is the ratio to virus only treatment. **(B-C)** Affinity of anti-E antibodies towards FcγIIa and FcγIIIa IgG receptor for individuals receiving different vaccine platforms. Fold change is measured against IgG isotype unspecific for Dengue. * p<0.05, ** p<0.01, het: heterology.
